# High Initial Heart Rate Score is an Independent Predictor of New Atrial High-Rate Episodes in Pacemaker Patients with Sinus Node Dysfunction

**DOI:** 10.1101/2024.02.07.24302470

**Authors:** Katsuhide Hayashi, Haruhiko Abe, Brian Olshansky, Arjun D. Sharma, Paul W. Jones, Nicholas Wold, David Perschbacher, Ritsuko Kohno, Gregory Y. H. Lip, Bruce L. Wilkoff

## Abstract

**Background:** Heart Rate Score (HRSc), the percent of atrial beats in the largest paced/sensed 10-bpm histogram bin recorded in cardiac devices, is a metric of chronotropic incompetence. It remains uncertain if HRSc independently predicts atrial high-rate episodes (AHREs) in patients with sinus node dysfunction (SND) undergoing pacemaker (PM) implantation.

**Objective:** To determine the relationship between initial HRSc post-PM implant and new-onset AHREs in patients with SND.

**Methods:** The cohort included patients with Boston Scientific PMs implanted for SND from 2012-2021 at Cleveland Clinic, University of Occupational and Environmental Health, Japan, Kyushu Rosai Hospital, and JCHO Kyushu Hospital. Patients were excluded if they had atrial fibrillation before PM implant or AHREs in the initial 3-months post-implant. Subsequent AHREs post-implant (>1% of atrial beats>170 bpm) were evaluated.

**Results:** Over 48.9 (IQR 25.8-50.4) months, 136 consecutive PM patients (age 75±10 years, 42% male) were followed. The median initial HRSc was 73(55-86)%. AHREs developed in 28/136 (21%). Although %RA pacing and HRSc correlated, rate-responsive pacing was associated with a HRSc only in the patients with high %RA pacing. Patients with HRSc≥80% had higher occurrence of AHREs than those with HRSc<80% (p=0.01, log-rank test). After adjusting for age, race, comorbidities, left ventricular ejection fraction, left atrial diameter, %RA/RV pacing, and rate-response programming, HRSc (HR:2.84, 95% CI:1.17-6.92; P=0.02) and male sex (HR:2.55, 95% CI:1.15-6.19; P=0.04) were independent predictors of AHREs.

**Conclusion:** HRSc≥80% independently predicted new-onset, device-determined AHREs for patients undergoing PM implant for SND. HRSc may have prognostic and therapeutic implications.

## Introduction

Heart Rate Score (HRSc) is a novel parameter of chronotropic incompetence using long-term heart rate variation for patients with cardiac implantable electronic devices (CIEDs).^1^ It is defined as the percent of atrial beats in the largest paced/sensed 10-beat/minutes histogram bin of a pacemaker (PM) or implantable cardioverter-defibrillator (ICD) (Figure 1). HRSc ≥70% predicts mortality in patients without atrial fibrillation (AF) who have ICDs^2^ or cardiac resynchronization therapy defibrillators (CRT-Ds).^3,4^

**Figure 1.**
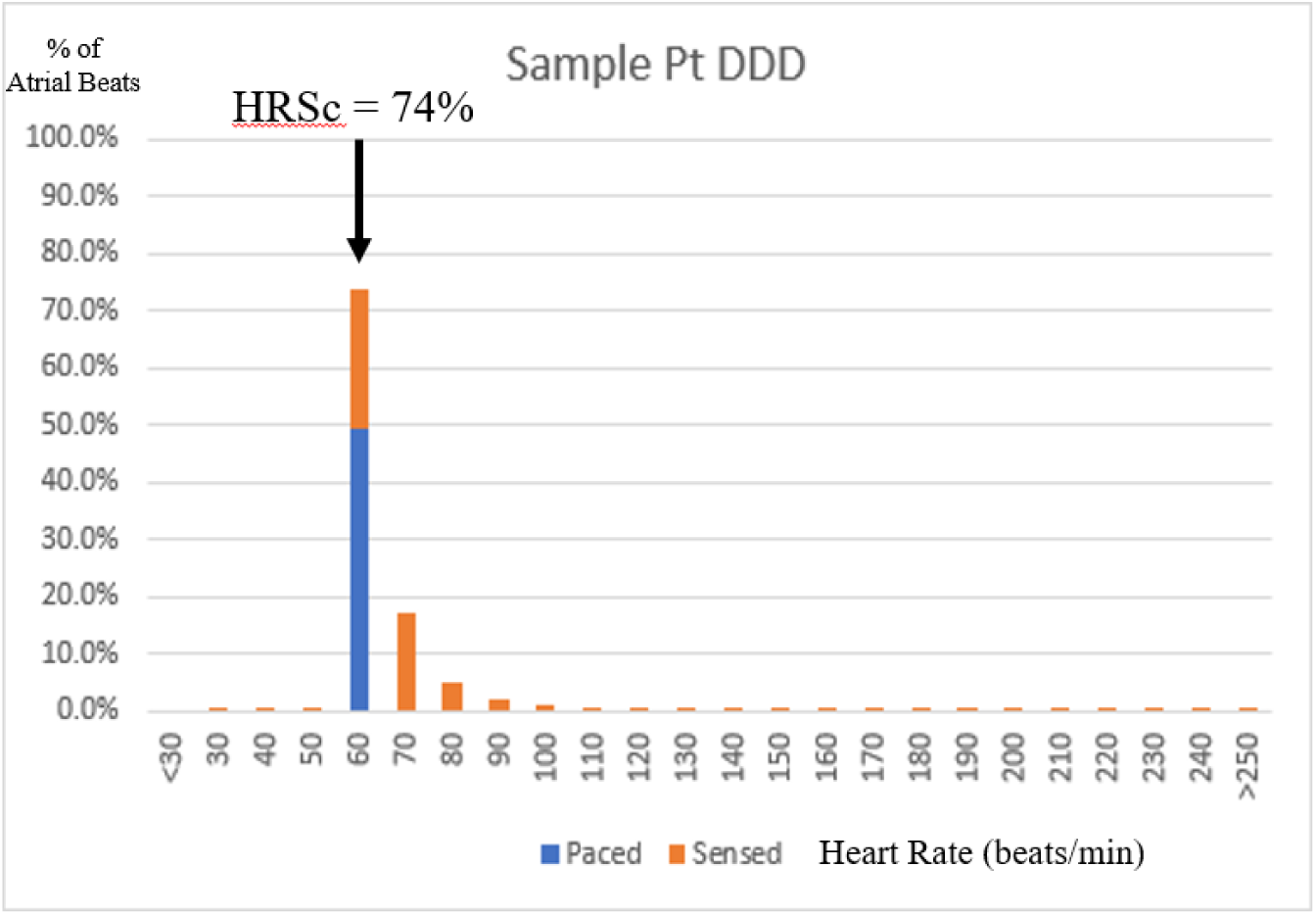
Example of HRSc. The HRSc was defined as the percent of all paced and sensed atrial event in single most populated 10-bpm rate histogram bin. This figure shows an example of HRSc of the patients in DDD mode. Here, HRSc is calculated as 74%. HRSc indicates heart rate score.

Atrial high-rate episodes (AHREs) have attracted much attention in anticoagulant therapy decisions for the risk of stroke in CIED patients.^5,6^ Unfortunately, no established data from electrocardiograms or device interrogation can yet predict development of AHREs. We recently showed that HRSc, early post-implantation, can predict new-onset AHREs in PM patients in a large database.^7^ In this prior study, the incidence of AHREs, defined as >1% of atrial beats ≥170 bpm (primary endpoint), correlated with the incidence of mode switch episodes ≥24 hours. However, that study could not collect clinical information, including PM indication, as it used a retrospective, de-identified, database. Therefore, it remains uncertain whether HRSc predicts AHREs in PM patients with sinus node dysfunction (SND) independent of clinical parameters.

The goal of this study was to evaluate the baseline HRSc post-PM implant in lieu of clinical characteristics and determine if an independent relationship exists between the initial HRSc and subsequent development of AHREs in patients with SND who underwent dual-chamber PM implant. We focused on the incidence of AHREs developing after implant for those with no AHREs during an initial 3-month observation.

## Methods

### Study Cohort

The cohort included consecutive patients with Boston Scientific PMs implanted for SND from 2012-2021 at 4 hospitals (Cleveland Clinic in the Unites States, University of Occupational and Environmental Health, Japan, Kyushu Rosai Hospital, and JCHO Kyushu Hospital in Japan). Patients were excluded if they had single chamber or biventricular PMs, atrial fibrillation (AF) before PM implantation or detected device-determined AHREs detection during the first 3 months after implantation.

Data were analyzed retrospectively. The study was approved by the Institutional Review Board of the Cleveland Clinic, University of Occupational and Environmental Health, Japan, Kyushu Rosai Hospital, and JCHO Kyushu Hospital. Heart Rate Score (HRSc) and Baseline Period HRSc was defined, as previously described, as the percent of all paced and sensed atrial events in the most populated 10-bpm rate histogram bin of the CIED, shown in Figure 1.^4^ HRSc, used in all analyses, was determined from remote interrogation data from the dual-chamber PM during the baseline period (the first 3 months after PM implantation). The HRSc, calculated at the last transmission during the baseline period, was designated the initial HRSc.

### Outcome

The endpoint was device-determined AHREs after the baseline period. The endpoint for AHREs was considered >1% of atrial beats ≥170 beats/min recorded by the PM.

### Follow-up Data Analysis

The occurrence of AHREs was examined using stored remote monitoring data and tracked during follow-up periods. The relationship between the initial HRSc and occurrence of device-determined AHREs was assessed.

### Statistical Analysis

Continuous variables were expressed as mean±standard deviation (SD), median (interquartile range, IQR: 25^th^-75^th^), or range, depending on the distribution of data, whereas categorical variables were expressed as counts and percentages. The optimal HRSc for predicting device-determined AHREs was chosen to be 80% for AHREs based on J-statistics (Figure 2). Kaplan-Meier analyses evaluated freedom from AHREs after the baseline period for the two initial HRSc groups. Log-rank tests were performed to compare group classified by initial HRSc. Variables with P values <0.05 after single variable analysis were entered into a multiple variable regression analysis to assess if there is an independent predictor of AHREs. P<0.05 was considered statistically significant. All statistical analysis were performed using JMP version 15.2.0 (SAS Institute Inc., Cary, NC).

**Figure 2.**
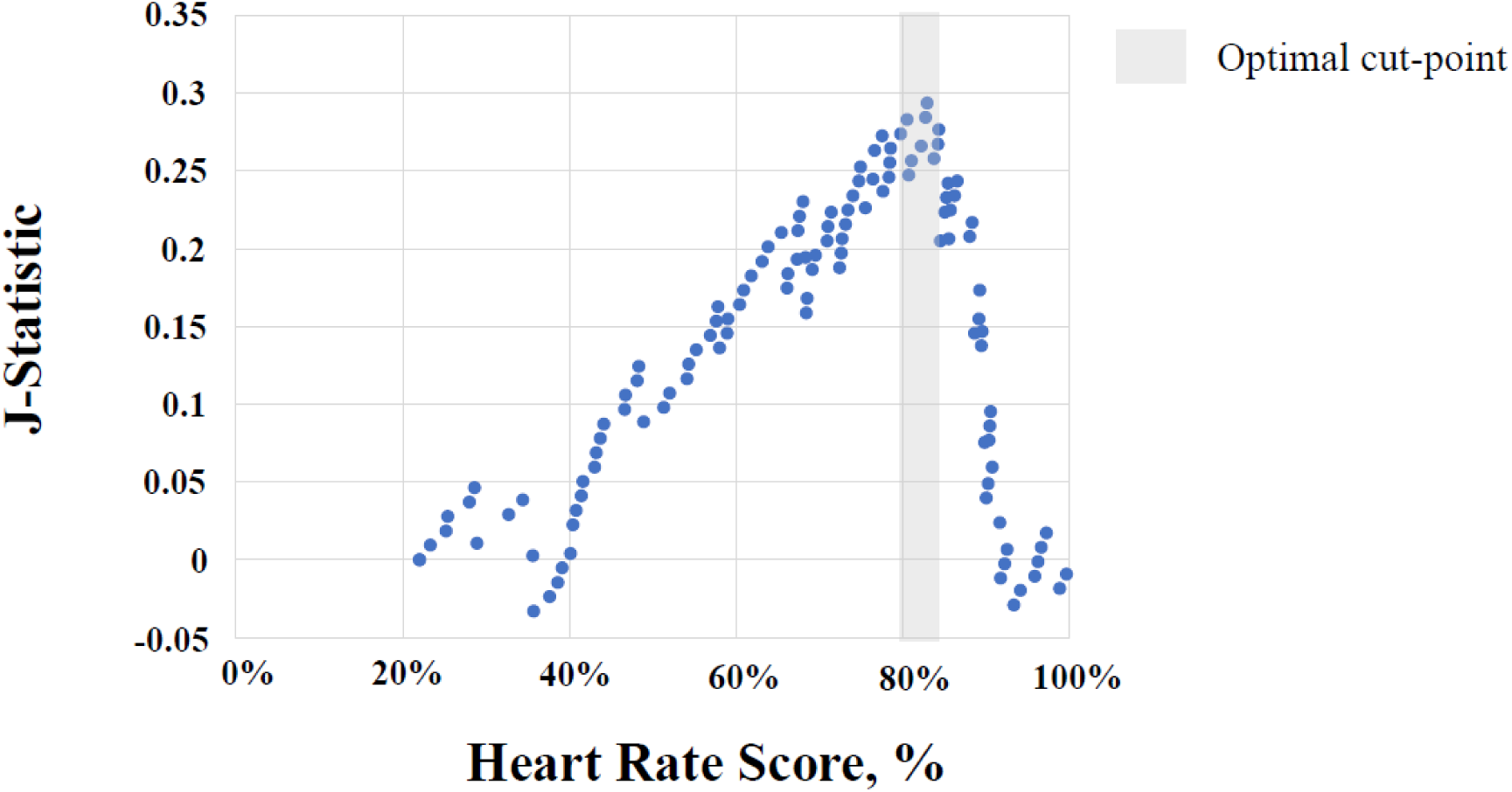
Optimal HRSc for predicting AHREs. J-statistic analysis shows 80% as the optimal HRSc for predicting device-determined AHREs. AHREs, atrial high-rate episodes; HRSc, heart rate score.

## Results

### Subjects Demographics

A total of 310 consecutive patients were enrolled. Patients who had atrial fibrillation before PM implantation or device-determined AHREs in the first three months (n=174) were excluded. The remaining 136 were followed for a median of 48.9 (IQR, 25.8-50.4) months.

Patient characteristics are listed in Table 1. The mean patient age was 75±10 years; 57 (41.9%) were male. Almost 50% of patients were Asian and 40% were European American. No patients were prescribed anti-arrhythmic drugs. The mean left ventricular ejection fraction (LVEF) and left atrial diameter (LAD) by echocardiogram were 62±7 % and 41±7 mm, respectively. The atrial lead was placed in the right atrium appendage in almost all patients. The ventricular lead was implanted in right ventricular septum in approximately two-thirds.

**Table 1.**
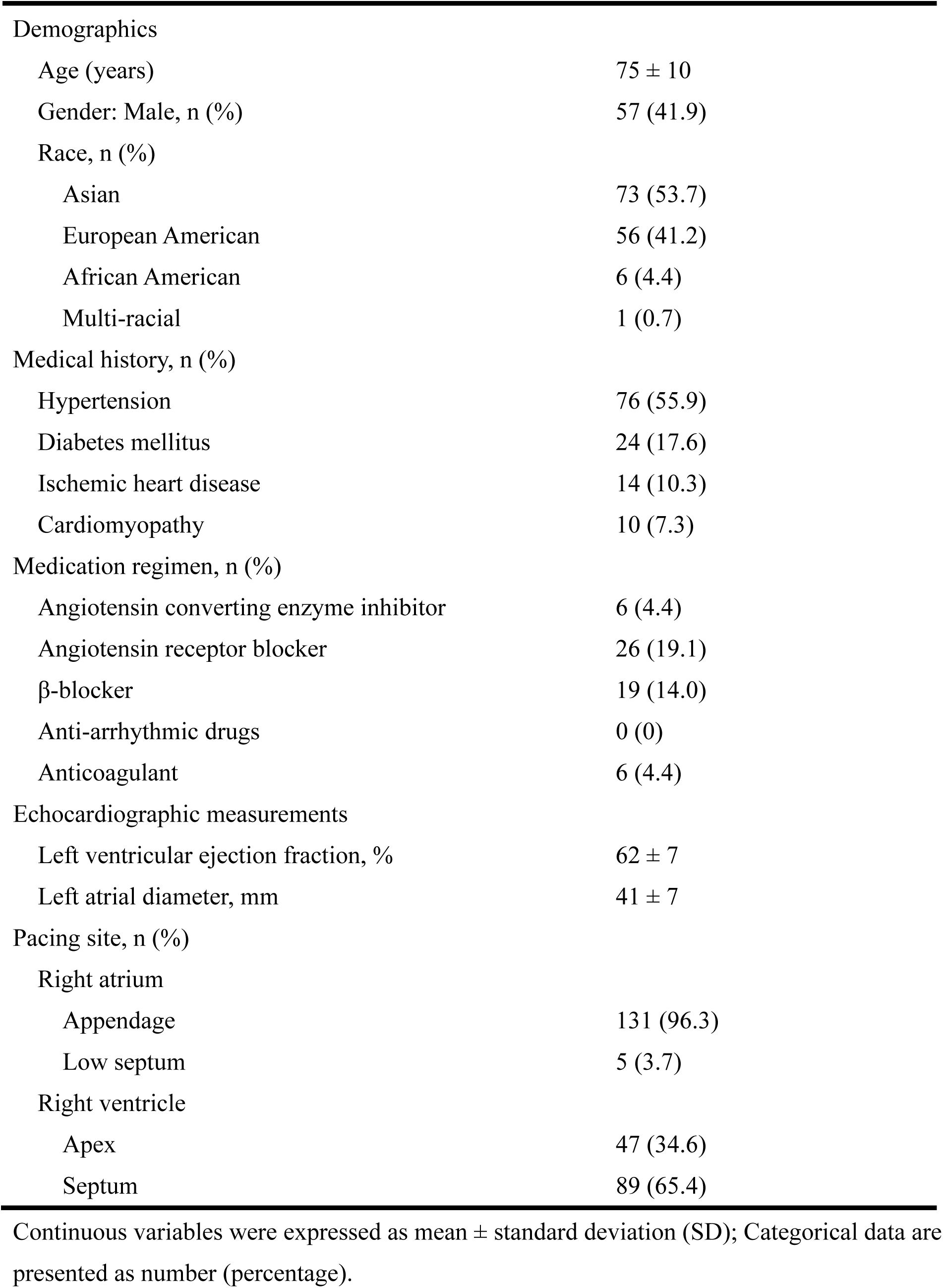
Patient characteristics.

Demographics
|  |  |
| --- | --- |
| Age (years) | 75 ± 10 |
| Gender: Male, n (%) | 57 (41.9) |
| Race, n (%) |  |
| Asian | 73 (53.7) |
| European American | 56 (41.2) |
| African American | 6 (4.4) |
| Multi-racial | 1 (0.7) |
| Medical history, n (%) |  |
| Hypertension | 76 (55.9) |
| Diabetes mellitus | 24 (17.6) |
| Ischemic heart disease | 14 (10.3) |
| Cardiomyopathy | 10 (7.3) |
| Medication regimen, n (%) |  |
| Angiotensin converting enzyme inhibitor | 6 (4.4) |
| Angiotensin receptor blocker | 26 (19.1) |
| β-blocker | 19 (14.0) |
| Anti-arrhythmic drugs | 0 (0) |
| Anticoagulant | 6 (4.4) |
| Echocardiographic measurements |  |
| Left ventricular ejection fraction, % | 62 ± 7 |
| Left atrial diameter, mm | 41 ± 7 |
| Pacing site, n (%) |  |
| Right atrium |  |
| Appendage | 131 (96.3) |
| Low septum | 5 (3.7) |
| Right ventricle |  |
| Apex | 47 (34.6) |
| Septum | 89 (65.4) |

2 Continuous variables were expressed as mean ± standard deviation (SD); Categorical data are 3 presented as number (percentage).

### Pacemaker Programming and Interrogation Data

Pacemaker programming at the time of implantation is shown in Table 2.

**Table 2.**
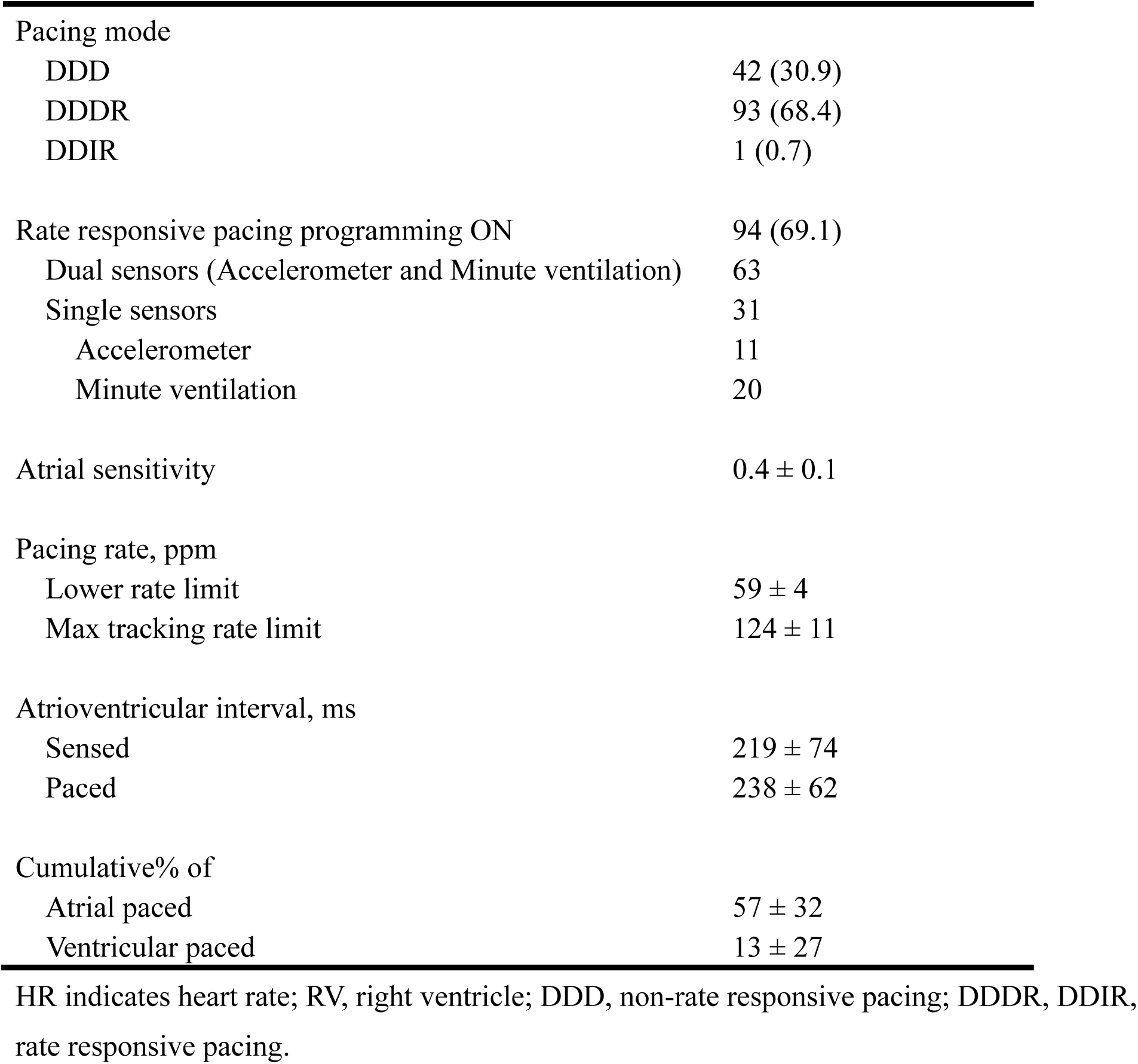
Pacemaker programming and interrogation data.

Rate-responsive pacing was programmed in 70% of patients. Atrial sensitivity was programmed to a mean of 0.4±0.1 mV and the lower rate limit (LRL) was programmed to 60 bpm in 124/136 (91.2%) patients. The cumulative percent of RA and RV pacing (%RA pacing and %RV pacing) during the baseline period were a mean of 57±32 %, and 13±27 %, respectively.

### Heart Rate Score

The initial HRSc was calculated at a median of 51 days (IQR, 7-84) after pacemaker implantation. The median value of the HRSc was 73% (IQR, 55-86%).

### Outcome

Over a median of 48.9 months (IQR, 25.8-50.4 months), 28/136 (21%) subjects experienced the endpoint of >1% AHREs. Subjects with an initial HRSc ≥80% had a higher incidence of AHREs (Kaplan-Meier rate of 40%) versus those with an initial HRSc <80% (Kaplan-Meier rate of 24%) through the 90.0 months during post-baseline follow-up. (log-rank P =0.01) (Figure 3).

**Figure 3.**
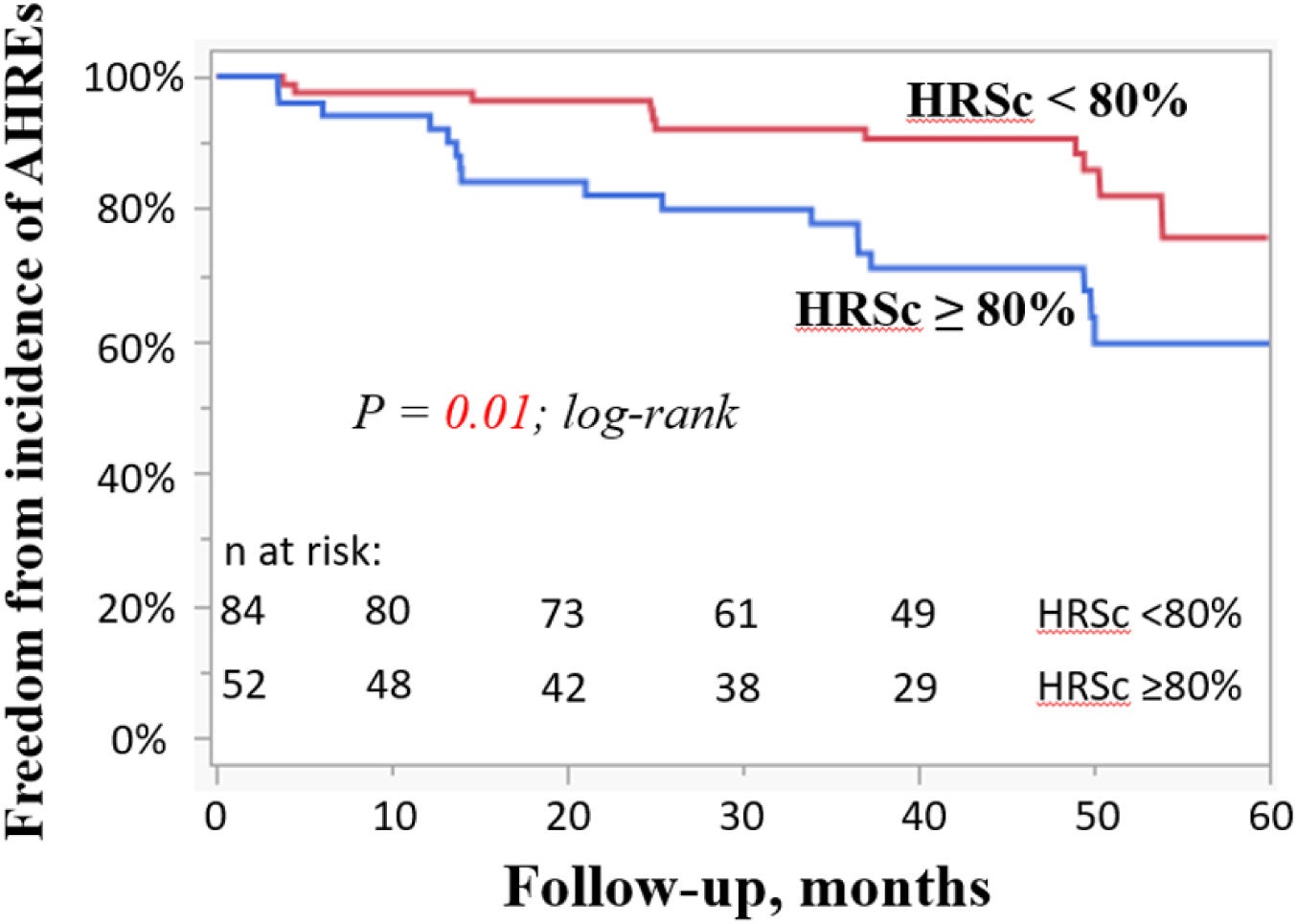
Survival free from AHREs by HRSc. Survival free from AHREs by HRSc Grouping is shown in this Kaplan-Meyer analysis. Kaplan-Meier analysis shows that subjects with an initial HRSc ≥80% had a higher incidence of AHREs versus those with an initial HRSc <80%, the primary end point. HRSc indicates heart rate score; AHREs, atrial high-rate episodes.

### Association of Heart Rate Score and Percent of Right Atrial Pacing

Figure 4 shows a positive correlation between %RA pacing during the baseline period and initial HRSc (R^2^=0.725, P<0.0001) for all patients. Patients were analyzed additionally by the %RA pacing and rate-response (ON or OFF). HRSc was plotted against RA pacing (Figures 5 and 6). In all 7 patients with high %RA pacing (pacing ≥70%) and no rate response (OFF), HRSc was high (≥80%). For those with rate response ON, with RA pacing ≥70%, 14 /54 patients had HRSc <80% (Figure 5). In patients with mid and low %RA pacing (<70%), the distribution of HRSc did not differ between those with rate response ON and OFF (Figure 6).

**Figure 4.**
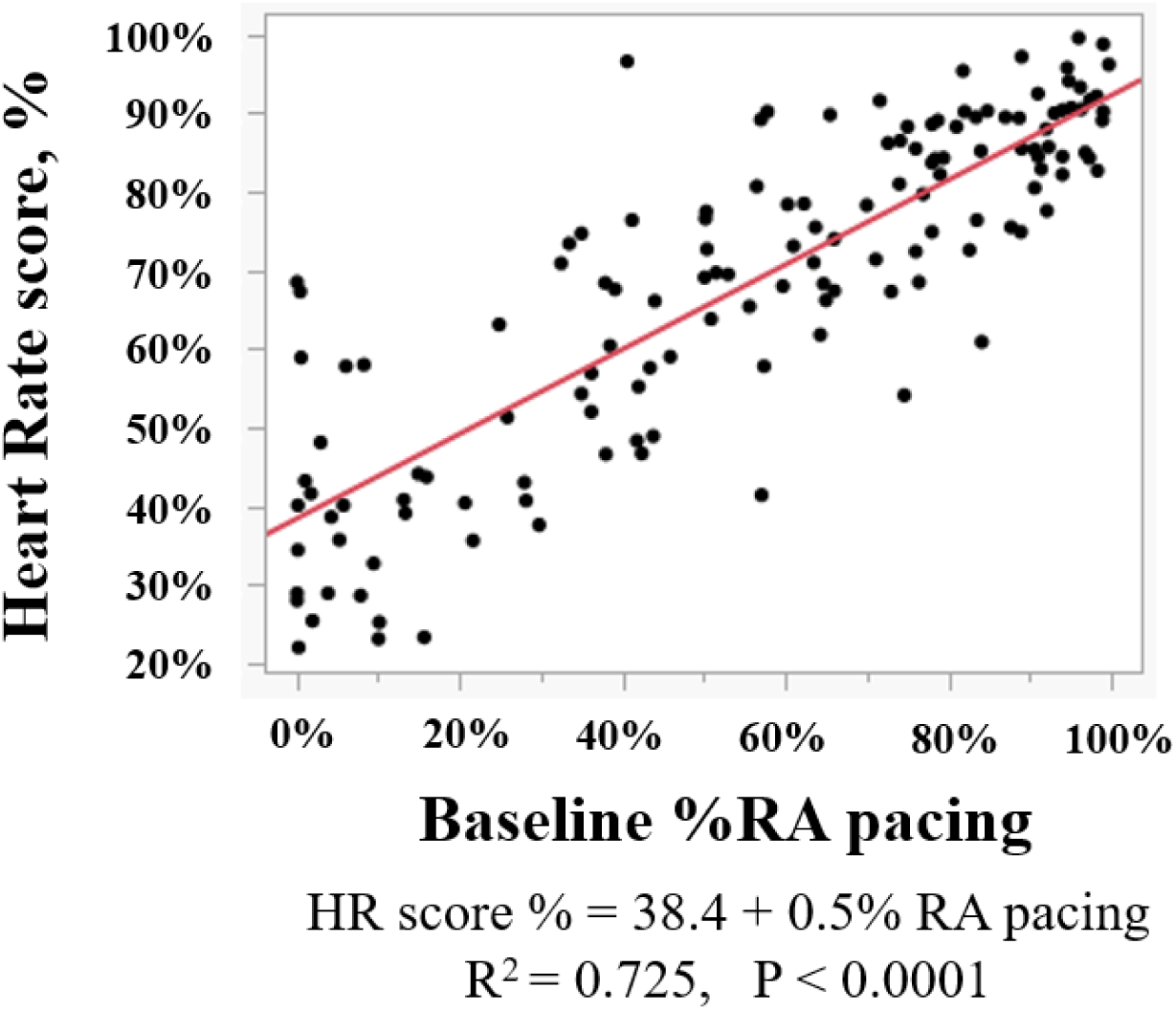
Heart Rate Score and % RA pacing (Correlation by continuous measures) The %RA pacing and initial HRSc were strongly correlated.

**Figure 5.**
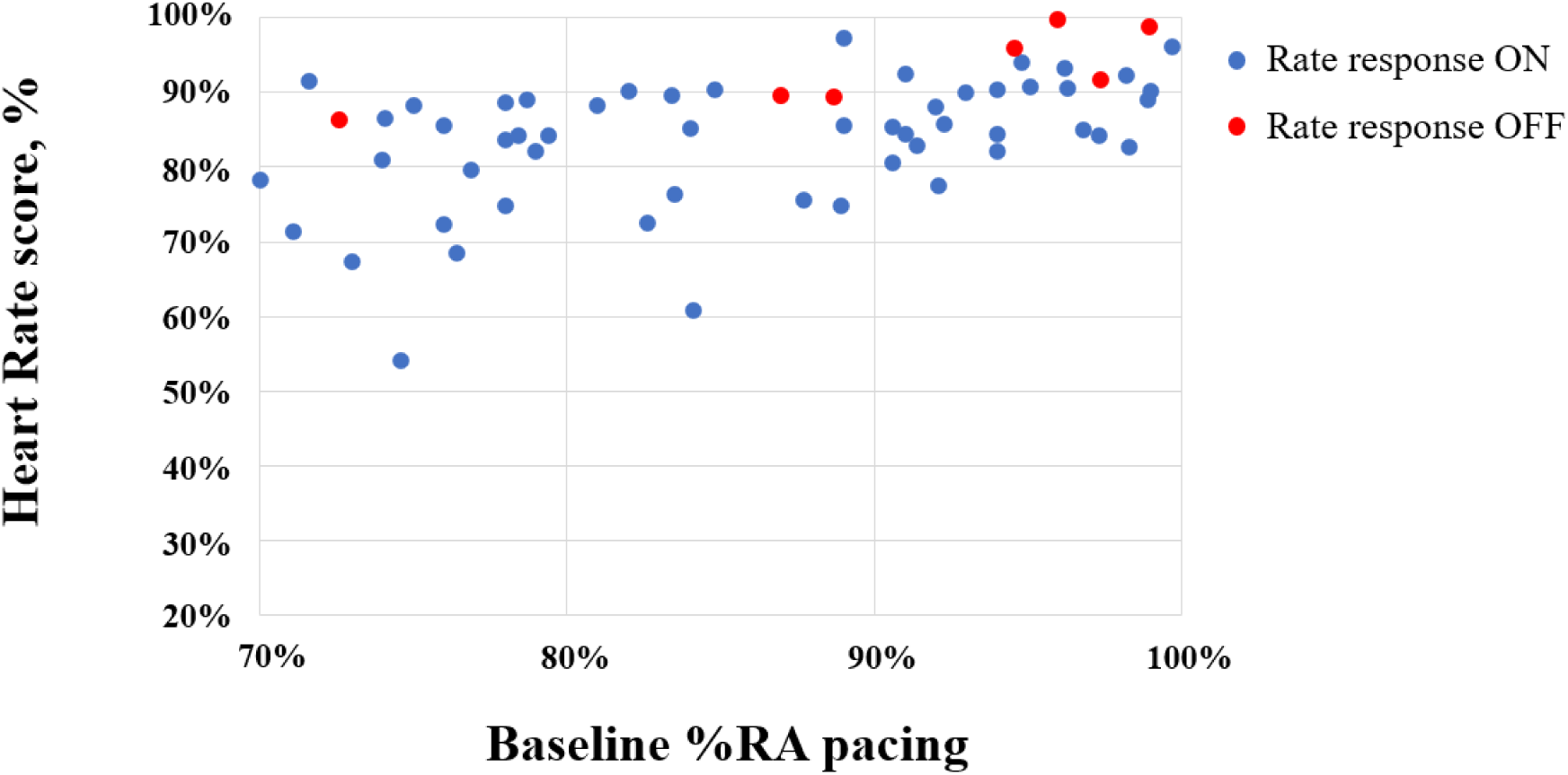
Heart rate score and %RA pacing (%RA pacing ≥70%, N=61) In the patients with %RA pacing ≥70%, when the rate response was OFF (red point, N=7), all HRSc was ≥80%. On the other hand, when the rate response was ON (blue point, N=54), 14/54 patients had HRSc <80%

**Figure 6.**
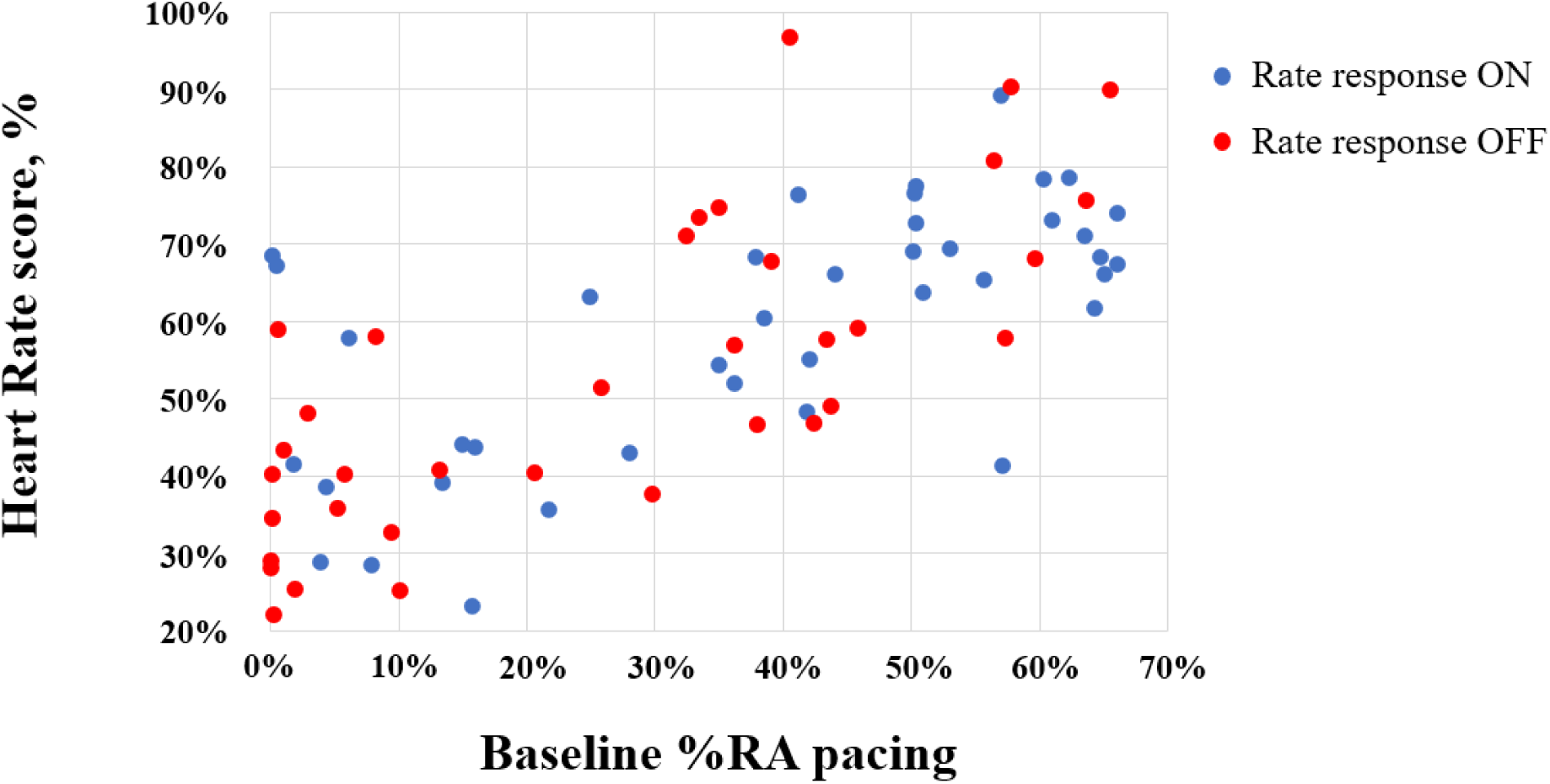
Heart rate score and %RA pacing (%RA pacing <70%, N=75) In the patients with RA pacing <70%, the distribution of HRSc did not differ between the cases with rate response ON (blue point, N=40) and OFF (red point, N=35). HRSc indicates heart rate score; RA, right atrium.

### Predictors for Incidence of Atrial High-Rate Episode

By multivariable analysis, the initial HRSc (HR: 2.84, 95% CI: 1.17-6.92; P=0.02) and male sex (HR: 2.55, 95% CI: 1.15-6.19; P=0.04) were the only independent predictors of AHREs incidence (Table 3).

**Table 3.**
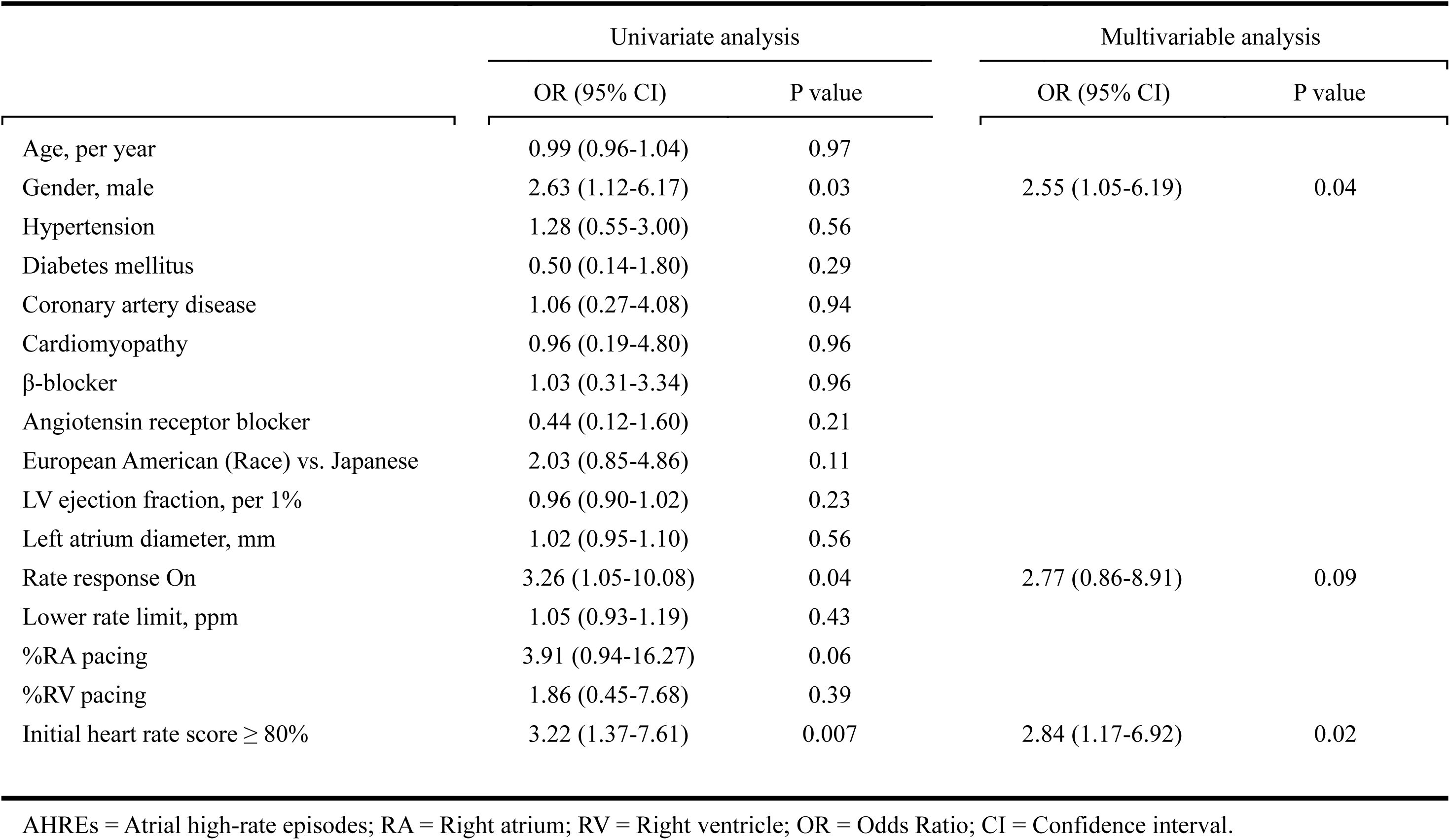
Predictors for AHREs.

## Discussion

We found that the initial HRSc ≥80 %, calculated within 3 months after dual-chamber PM implantation for SND, independently predicted new-onset, device defined, AHREs in subsequent follow-up. This report is the first to demonstrate a relationship between HRSc and device-detected AHREs in this population. Subjects with an initial HRSc ≥80% had an increased risk of AHREs. This analysis held true over the 48.9-month follow-up, after adjustment for risk factors for AHREs including: age, race, histories of hypertension, diabetes mellitus, coronary artery disease, cardiomyopathy, LVEF, LAD, %RA/RV pacing, and rate-responsive pacing. Despite a positive correlation between %RA pacing and HRSc for patients dependent on RA pacing, more than one-quarter with rate-responsive pacing had HRSc <80%; those without rate-responsive pacing all had HRSc ≥80%.

Our study indicates that a high initial HRSc (≥80%) predicts AHREs recorded after PM implantation for SND independently. Currently no established indicator from electrocardiograms or device interrogation has been shown to predict subsequent development of AHREs. Although our prior report^7^ indicated that the HRSc predicts subsequent AHREs in PM patients in a large de-identified database, clinical parameters including PM indication could not be determined. Therefore, we investigated the relationship between initial HRSc and subsequent AHREs focusing on PM patients implanted for SND in the current study.

Our results demonstrate that HRSc predicts pacemaker detected AHREs. It is not clear that this correlates directly to clinical AF. However, in our prior report^7^, the incidence of AHREs defined by >1% of atrial beats ≥ 170 bpm correlated with the incidence of mode switch episodes ≥24 hours which is likely to represent substantial paroxysmal or persistent AF.

Age, gender, hypertension, diabetes mellitus, ischemic heart disease, pharmacologic therapy, left atrial dilatation, and LVEF, are well known risk factors for AF.^8–11^ In our study, HRSc predicted AHREs independent of other risk factors. Interestingly, however, male gender also predicted AHREs. This result is consistent with community-based ARIC (Atherosclerosis Risk In Communities) study that showed higher incidence of AF in males than females.^12^ In pacemaker patients with SND, high %RV pacing is found to be associated with an increased AF.^13–15^ In addition, the association between %RA pacing and AF has been also identified.^16,17^

In the current study, the median HRSc of 73% in PM patients with SND was higher than prior reports.^2,7^ This result indicates that our study patients have severe chronotropic incompetence. Patients with little or no spontaneous heart rate change above the LRL have a high HRSc^1^. In addition, the optimal HRSc cut-point to predict device-determined AHREs was found to be ≥80%. Notably, this optimal cut-point is different from HRSc ≥70% for predicting mortality in patients with ICDs or CRT-Ds.^3,4^

Rate-responsive pacing can decrease HRSc in patients with sinus node dysfunction who pace predominantly at the LRL and have a high HRSc (Figure 5). On the other hand, for patients with SND who have less chronotropic incompetence, rate-responsive RA pacing may be less likely affect HRSc (Figure 6). Rate responsive pacing may be excessive or even harmful for those who have chronotropic competence.^3^ Rate-responsive pacing may be best reserved for those with high baseline HRSc. However, LRL programming affects %RA pacing and HRSc. A high LRL can override the resting intrinsic rate resulting in an increase in %RA pacing and subsequently increase in HRSc. Conversely, programming a lower LRL may decrease %RA pacing and lower HRSc. Most subjects in this study were programmed to an LRL of 60 but prior data showed that lower LRL programming improved survival in a large database of CRT-D subjects.^18^ Further research is needed to assess the relationships of LRL, AHREs, %RA pacing and HRSc.

## Study Limitations

This was a retrospective analysis and therefore, may be affected by unknown confounders. We were unable to assess the impact of rate responsive pacing on AHREs. The present study also does not answer whether lowering HRSc with pacemaker programming affects AHREs. The population was not large enough to determine relationships of HRSc to a higher percentage of AHREs or episodes that could become persistent AF.

## Conclusion

In patients undergoing PM implant for SND, HRSc≥80% independently predicted device-determined AHREs in long-term follow-up. As initial HRSc predicts AHREs in this population, this parameter may have prognostic and therapeutic implications.

## Acknowledgements

Bruce L. Wilkoff, MD passed away before submitting this manuscript and authors express our sincerest condolence to him and sincerely appreciate his incredible leadership, he gave to see this project completed. And the authors sincerely appreciate Hiroyuki Takatsu, MD, of Kyushu Rosai Hospital, and Kan Kikuchi, MD, of JCHO Kyushu Hospital for obtaining patient’s demographic and HRSc data.

## Data Availability

The data underlying this article will be shared on reasonable request to the corresponding authors.

